# Healthcare workers’ knowledge, attitude and practices during the COVID-19 pandemic response in a tertiary care hospital of Nepal

**DOI:** 10.1101/2020.08.19.20177998

**Authors:** Dil K Limbu, Rano M Piryani, Avinash K Sunny

## Abstract

**Background:** To control the spread of ongoing COVID-19 infection, extremely important measures need to be adopted. Healthcare worker’s adherence to prevention and control measures is affected by their knowledge, attitudes, and practices (KAP) towards COVID-19. In this study, we assessed the KAP among healthcare workers towards COVID-19 during the ongoing pandemic.

**Method:** A self-developed piloted KAP questionnaire was used among the recruited healthcare workers working for the COVID-19 response in the Universal College of Medical Sciences Teaching Hospital (UCMSTH), in Bhairahawa, Nepal. The knowledge questionnaire consisted of questions regarding the clinical characteristics, prevention and management of COVID-19. Assessment on attitudes and practices towards COVID-19 included questions on behaviour and change in practices while working during this response. Knowledge scores were calculated and compared by demographic characteristics and their attitude and practices towards COVID-19. Data were analysed using bivariate statistics.

**Results:** A total of 103 healthcare workers participated in the study. The mean age of the participants was 28.24±6.11 years (range: 20-56); 60.2% were females; 61.2% were unmarried; 60.2% had medical degree and 39.8% were nursing staff. The mean knowledge score was 10.59±1.12 (range: 7-13) and it did not vary by demographic characteristics. Attitude was positive for 53.4% participants with a mean knowledge score of 10.35±1.19 and negative for 46.6% participants with a mean knowledge score of 10.88±0.98 (p = 0.02). Practice was good (≥3 score) for 81.5% participants with a mean knowledge score of 10.73±1.12 and practice was poor for 18.5% participants with a mean knowledge score of 10.46±1.13 (p = 0.24). The attitude of the participants improved with the increasing age of the participants (29.55±7.17, p = 0.02).

**Conclusion:** There is comparably better knowledge regarding COVID-19 among healthcare workers along with appropriate practices, however attitude was less optimistic with better knowledge but more optimistic with higher age of the healthcare workers. Hence, protective measures for healthcare workers in order to improve their attitude is necessary during the pandemic response.

## Introduction

COVID-19 is a disease caused by the SARS-CoV-2 virus, first identified in the city of Wuhan, in China’s Hubei province in December 2019 (1). COVID-19 was previously known as 2019 novel Corona virus (2019-nCoV) respiratory disease before the World Health Organization (WHO) declared the official name as COVID-19 in February 2020 (2). In March 11, 2020, the World Health Organization (WHO) declared the COVID-19 outbreak a pandemic (3). This ongoing pandemic has been spreading very rapidly with more than 8.5 million confirmed infections and more than 0.47 million deaths worldwide till June 22 2020 (GMT 01.18) (4).

Countries worldwide have used various control measures such as social distancing, hand washing, closing of public transportation and public places, testing and tracing affected community (5). Like many governments around the world, Nepal has also called for lockdown since 24^rd^ March, 2020, allowing only essential services to be available like hospitals, groceries and medical supplies and frontline emergency services (6). In Nepal, the total number of confirmed infections stood at 9561 till June 22, 2020 (7).

Profound knowledge supports optimistic attitude and appropriate practice at work which in due course helps deter the risk of infection (8). Healthcare worker’s adherence to control measures is affected by their knowledge, attitudes, and practices (KAP) towards COVID-19, hence it is important to understand their knowledge and determine the factors that affect their attitudes and practices in order to have adequate guidance and protection. Thus, this study aimed to assess the KAP among healthcare workers towards COVID-19 during the ongoing pandemic.

## Method

### Study design and setting

A cross-sectional study was conducted from April 7, 2020 to May 7, 2020 at Universal College of Medical Sciences Teaching Hospital (UCMSTH), Bhairahawa, Nepal which has recently stepped its efforts to support government in its efforts towards COVID 19. UCMSTH is a teaching hospital serving as a tertiary care centre located in the southern west border of Nepal. All the healthcare workers working in the hospital during the COVID-19 pandemic response were included in the study.

### Data collection and management

Data were collected from healthcare workers using a self-administered questionnaire to assess KAP towards COVID-19. This questionnaire consisted of two parts with demographic information and a self-developed piloted and pretested KAP questionnaire (2^nd^ part) assessing knowledge attitude and practice. Knowledge was measured through 13 questions: 5 regarding clinical manifestation, 2 on the mode of transmission, and 6 for prevention and control of COVID-19 with a true/false/don’t know option. The reliability analysis of knowledge questionnaire with Cronbach’s alpha coefficient was found to be acceptable with an internal consistency of 0.76. Attitude and practice were assessed through 5 questions each with a yes/no/don’t know option. (Table 1)

Data were then entered into MS Excel and coded for anonymity. The entered data were exported into Statistical Package for Social Sciences (SPSS) version 23 for analysis.

### Study variables

***Demographic variables*** include age as continuous variable; gender as male and female; marital status married and unmarried; education as certificate level with Auxiliary Nurse Midwife (ANM), Proficiency Certificate Level (PCL) Nursing and Community Medical Assistant (CMA); Bachelor in Nursing Science (BNS); graduate medical education with Bachelor of Medicine and Bachelor of Surgery (MBBS); and post-graduate medical education with Doctor Medicine (MD), Master in Dental Surgery (MDS) and Doctorate of Medicine (DM); and designation as Nursing staff, Medical Intern, Medical Officer, Post Graduate (PG) Resident and Consultant.

***Knowledge scores*** were calculated by assigning 1 point to each correct answer and an incorrect/unknown answer was assigned 0 points. The total knowledge score ranged from 0 to 13, with a higher score signifying a better knowledge.

***Attitude*** was assessed as positive and negative. The average score on attitude questionnaire was calculated and used as a cut off for positive and negative.

***Practice*** was assessed as good and poor. The average score on practice questionnaire was calculated and used as a cut off for good and poor.

### Statistical analysis

Descriptive statistics were calculated as frequency, percentage, mean and standard deviation (SD). Data were analysed using Pearson chi-square test, Pearson correlation, independent t-test and one-way analysis of variance (ANOVA) test. At 95% Confidence Interval, p-value < 0.05 was considered to be statistically significant.

### Ethical approval

Ethical clearance was obtained from Institutional Review Committee (IRC) of Universal College of Medical Sciences Teaching Hospital (UCMSTH). Informed written consent was taken from the participant before inclusion in the study and confidentiality was maintained.

## Result

The correct responses on KAP are mentioned in table 1. A total of 103 healthcare workers participated in the study with a male to female ratio of 1:1.5. The mean age of the participants was 28.24±6.11 years (range: 20-56), 62 (60.2%) were females, 63 (61.2%) were unmarried, 62 (60.2%) had medical degree and 41 (39.8%) were nursing staff. The mean knowledge score was 10.59±1.12 range: 7-13) with 81.5% correct answer rate. There was no significant correlation in the knowledge score of the participants and their mean age (p = 0.13). The mean knowledge score for male participants was 10.76±1.16 while for female participants 10.48±1.10, however the difference was not statistically significant (p = 0.24). Married participants had mean knowledge score of 10.78±1.00 while for unmarried participants it was 10.48±1.19, suggesting no statistically significant difference (p = 0.19). By education, the mean knowledge score was highest for MBBS education with 10.85±1.28 (p = 0.06) and by designation; it was highest for Resident with 11.00±0.86 (p = 0.05), however the difference was not statistically significant. (Table 2)

**Table 1.** Correct responses on KAP questionnaire (n = 103) Knowledge Questions.

| Knowledge Questions | Correct response (%) |
| --- | --- |
| 1. Fever, dry cough, difficulty in breathing, tiredness are the common clinical symptoms of COVID-19. | 103 (100%) |
| 2. Sneezing, runny nose, stuffy nose and headache are less common in persons infected with COVID-19. | 80 (77.7%) |
| 3. Loss of taste and smell are also the feature of COVID 19 infection. | 52 (50.5%) |
| 4. Currently there is no treatment of COVID-19 infection, but early symptomatic and supportive treatment can help most patient recover from infection. | 101 (98.1%) |
| 5. Majority of COVID-19 infective patient will not develop severe illness but elderly, patient having chronic illness, DM, COPD are likely to develop severe illness. | 86 (83.5%) |
| 6. COVID-19 infected person with fever can infect to other people. | 28 (27.2%) |
| 7. COVID-19 virus spread via respiratory droplets. | 99 (96.1%) |
| 8. Ordinary people should wear general mask. | 85 (82.5%) |
| 9. People maintain 2-meter distance in the public places. | 94 (91.3%) |
| 10. Lockdown is effective measure to slow the spread of infection. | 102 (99.0%) |
| 11. People infected with COVID-19 should immediately place in proper isolation. | 102 (99.0%) |
| 12. Suspected COVID19 patient should be sent to a quarantine centre or home quarantine. | 99 (96.1%) |
| 13. Health care professional with direct contact should take tablet hydroxychloroquine as a prophylaxis. | 60 (58.3%) |
| **Attitude Questions** | **Correct response (%)** |
| 1. Can Nepal win the battle against COVID-19? | 52 (50.5%) |
| 2. Are you confident to work in hospital during COVID-19 pandemic? | 47 (45.6%) |
| 3. Does your family support you to work in hospital during pandemic? | 58 (56.3%) |
| 4. Do you experience anxiety and fear while working with suspected COVID-19 patient? | 81 (78.6%) |
| 5. Have all the doctors from various department actively involved in COVID-19 Pandemic response? | 38 (36.9%) |
| **Practice Questions** | **Correct response (%)** |
| 1. Are you being trained to work for COVID-19 patient? | 18 (17.5%) |
| 2. Have you following social distancing? | 78 (75.5%) |
| 3. Have you been wearing mask and gloves during hospital practice? | 99 (96.1%) |
| 4. Do you regularly follow infection protection measures? | 87 (84.5%) |
| 5. Are you attending patient suspected with COVID-19? | 71 (68.9%) |

**Table 2.** Knowledge scores of COVID-19 by demographic variables (n = 103) Characteristics Category N (%) Knowledge score.

| Characteristics | Category | N (%) | Knowledge score (Mean ± SD) | p-value |
| --- | --- | --- | --- | --- |
| **Age (years)** | 28.24±6.11 | 103(100) | 10.59±1.12 | 0.13<sup>a</sup> |
| **Gender** | Male | 41(39.8) | 10.76±1.16 | 0.24<sup>b</sup> |
| | Female | 62(60.2) | 10.48±1.10 | |
| **Marital status** | Unmarried | 63(61.2) | 10.48±1.19 | 0.19<sup>b</sup> |
| | Married | 40(38.8) | 10.78±1.00 | |
| **Education** | ANM/PCL/CMA | 34(33.0) | 10.18±1.03 | 0.06<sup>c</sup> |
| | BNS | 7(6.8) | 10.57±0.96 | |
| | MBBS | 47(45.6) | 10.85±1.28 | |
| | MD/MDS/DM | 15(14.6) | 10.73±1.12 | |
| **Designation** | Nursing staff | 41(39.8) | 10.24±1.18 | 0.05<sup>c</sup> |
| | Medical Intern | 23(22.3) | 10.87±0.92 | |
| | Medical Officer | 4(3.9) | 10.00±1.41 | |
| | Resident | 20(19.4) | 11.00±0.86 | |
| | Consultant | 15(14.6) | 10.73±1.28 | |
| **Attitude** | Positive ($\geq 3$) | 55(53.4) | 10.35±1.19 | **0.02<sup>d</sup>** |
| | Negative ($< 3$) | 48(46.6) | 10.88±0.98 | |
| **Practice** | Good ($\geq 3$) | 84(81.5) | 10.73±1.12 | 0.24<sup>d</sup> |
| | Poor ($< 3$) | 19(18.5) | 10.46±1.13 | |

<sup>a</sup>Pearson correlation, <sup>b</sup>Independent t-test, <sup>c</sup>one way ANOVA test, <sup>d</sup>Pearson chi-square test

The mean knowledge score varied significantly with the attitude of the participants (p = 0.02) but it did not vary with their practice (p = 0.24). Attitude was positive (≥3 score) for 55(53.4%) participants with a mean knowledge score of 10.35±1.19 and negative (≤3 score) for 48(46.6%) participants with a mean knowledge score of 10.88±0.98. Similarly, practice was good (≥3 score) for 84(81.5%) participants with a mean knowledge score of 10.73±1.12 and practice was poor (< 3 score) for 19(18.5%) participants with a mean knowledge score of 10.46±1.13. There was negative correlation between knowledge and attitude (r = –0.313, p = 0.001), however knowledge didn’t correlate with practice (r = 0.093, p = 0.35). There was significant correlation between attitude and practice (r = 0.298, p = 0.002). (Table 2 & Table 3)

**Table 3.** Correlation between the Knowledge, attitude and practice scores (n = 103)

| **Variable**<br>**(Mean ± SD)** |  | **Knowledge** | **Attitude** | **Practice** |
| --- | --- | --- | --- | --- |
| **Knowledge**<br>(10.59±1.12) | **Correlation Coefficient (r)** | 1 | -0.313 | 0.093 |
|  | **p-value** |  | **0.001\*** | 0.350* |
| **Attitude**<br>(2.68±1.16) | **Correlation Coefficient (r)** | -0.313 | 1 | 0.298 |
|  | **p-value** | **0.001\*** |  | **0.002\*** |
| **Practice**<br>(3.43±1.03) | **Correlation Coefficient (r)** | 0.093 | 0.298 | 1 |
|  | **p-value** | 0.350* | **0.002\*** |  |

\* Pearson correlation

Association of attitude and practice of the participants towards COVID-19 with their characteristics were assessed. There was no significant difference in the attitude of the participants by their gender (p = 0.66), marital status (p = 0.51), education (p = 0.42) or designation (p = 0.28), however the attitude varied significantly with the increasing age of the participants (p = 0.02). The practice of the participants was mostly good and did not differ by age (p = 0.64), gender (p = 0.82), marital status (p = 0.47), education (p = 0.78) or designation (p = 0.09). (Table 4)

**Table 4.** Attitude and Practice towards COVID-19 by demographic variables (n = 103) Characteristics Category Positive attitude N.

| **Characteristics** | **Category** | **Positive**<br>**attitude N**<br>**(%)** | **p-value** | **Good**<br>**Practice N**<br>**(%)** | **p-value** |
| --- | --- | --- | --- | --- | --- |
| **Age (years)** |  | 29.55±7.17 | **0.02\*** | 28.38±6.10 | 0.64* |
| **Gender** | Male | 23(41.8) | 0.66 | 33(39.3) | 0.82 |
|  | Female | 32(58.2) |  | 51(60.7) |  |
| **Marital status** | Unmarried | 32(58.2) | 0.51 | 50(59.5) | 0.47 |
|  | Married | 23(41.8) |  | 34(40.5) |  |
| **Education** | ANM/PCL/CMA | 18(32.7) | 0.42 | 26(31.0) | 0.78 |
|  | BNS | 2(3.6) |  | 6(7.1) |  |
|  | MBBS | 25(45.5) |  | 40(47.6) |  |
|  | MD/MDS/DM | 10(18.2) |  | 12(14.3) |  |
| **Designation** | Nursing | 20(36.4) | 0.28 | 32(38.1) | 0.09 |
|  | Medical Intern | 9(16.4) |  | 16(19.0) |  |
|  | Medical Officer | 3(5.5) |  | 4(4.8) |  |
|  | Resident | 13(23.6) |  | 20(23.8) |  |
|  | Consultant | 10(18.2) |  | 12(14.3) |  |

\*independent t-test

## Discussion

This is a cross-sectional study conducted with objective to assess the KAP among healthcare workers towards COVID-19 during the ongoing pandemic The findings of this study related to overall correct answer (81.5%) are comparable with a similar study conducted in China which reported that 89% healthcare workers surveyed demonstrated sufficient knowledge on COVID-19 (9). As this study was conducted during the national lockdown period in Nepal, healthcare workers were quite aware of on most of the information related COVID-19 as part of being prepared for the response to the ongoing pandemic. Despite the differences in demographic characteristics of healthcare workers, the knowledge was better among all of the them. This is in contrast with the studies suggesting differences in knowledge by the type of healthcare workers (10).

Overall, 53.4% of the healthcare workers had positive attitude towards COVID-19. This finding is lower compared to that in other studies conducted in China. This speculation could probably due to as only a half of them (50.5%) believed that Nepal could win the battle against COVID-19 and despite having family support (56.3%), they were less confident (45.6%) while at work because of anxiety and fear (78.6%) working with suspected patients and not all the doctors (36.9%) from departments were actively involved in COVID-19 pandemic response. The attitude of the healthcare workers was found to be similar across their different demographic characteristics. This suggests that the notion was same for all in this pandemic situation, defying the reports from other studies of varying attitude by the healthcare workers demographics (11).

Better knowledge of COVID-19 among the Healthcare workers did not correlate with their attitude towards the disease while at work in this study. This finding is in contrast with other studies reporting that knowledge directly affected their attitude and increased their confidence (9,10). Knowledge is a prerequisite not only for establishing prevention beliefs and promoting positive behaviours but also for forming positive attitudes towards disease for better effectiveness of their coping strategies and behaviours to a certain extent (8). However, studies have reported that protective measures not being in place could increase the chances of infection while well protected emergency and other departments in the hospital had lower chances of infection (12). Healthcare workers with higher age elicited positive attitude in this study. Similar was the case among healthcare workers in other studies. Higher the age, longer is the experience in dealing with emergencies ultimately demonstrating confidence and optimism. Hence, increasing age could be the reason for positive attitude among the healthcare workers (13).

In this study, 81.5% of the healthcare workers practices was found to be appropriate and it did not differ by their demographic characteristics or knowledge scores. However, the practice significantly correlated with their attitude. Thus, poor practices can be linked to poor attitude and it resulted so because very few (17.5%) were trained to work for COVID-19 patient, despite many following practices such as social distancing (75.5%), wearing mask and gloves during hospital practice (96.1%), infection protection measures (84.5%) and attending patient suspected with COVID-19 (68.9%). This finding is comparable to a similar study in China which found 89.7% of the healthcare workers followed correct practices regarding COVID-19 (9). Moreover, the healthcare workers cannot neglect their own protection at work as they are the most vulnerable to infection (14).

## Limitations

This study has some limitations. The findings of this study should be cautiously used for generalization since it depicts one hospital of Nepal. Additionally, KAP assessment may be underestimated, hence further study is warranted.

## Conclusion

This study found out that there is better knowledge regarding COVID-19 among healthcare workers along with appropriate practices, however, attitude was less optimistic with better knowledge and more optimistic with higher age of the healthcare workers. Healthcare workers practice is directly correlated with their attitude. Hence, despite better knowledge, there is need for more positive attitude and better practices to be in place. Thus, education and training on protection and protective measures are required for subsequently improving positive attitude and better practices at work during the COVID-19 pandemic response.

## Data Availability

Yes - all data are fully available without restriction. All relevant data are within the manuscript and its Supporting Information files.

## Acknowledgements

We thank all the healthcare workers for their voluntary participation in the study.

